# Concerns over spillover, exclusions, and hospital unit differences in an AI early warning system trial (CONCERN-EWS)

**DOI:** 10.64898/2026.11.23.26368768

**Authors:** Sean Mann, Carl T. Berdahl, Beth Ann Griffin

**Author notes:** corresponding author Sean Mann. RAND, Arlington, VA, USA.

## Abstract

Rossetti et al. report that hospitalized patients whose care was informed by an AI-based clinical deterioration early warning system (EWS) were 36% less likely to die during a pragmatic cluster-randomized trial when compared to patients on usual care units without the EWS (adjusted hazard ratio [HR], 0.64; 95% confidence interval [CI], 0.53–0.78).^1^ The authors should be applauded for conducting a trial evaluating the effect of an AI intervention on patient health outcomes; more such trials are needed. Several aspects of the study, however, raise concerns that its findings may not accurately reflect the effect of the EWS. First, spillover effects on availability of scarce intensive care unit (ICU) beds could have led to worse outcomes for patients in the control group. Second, EWS-informed clinical decisions may have led to more frequent exclusion of high-risk patients from the intervention group. Either of these concerns could have led to overestimation of AI benefit. Finally, large pre-trial differences in hospital unit mortality rates suggest that unreported cluster-level imbalance or between-cluster variance could also have affected trial results.

## Main Text

Rossetti et al. report that hospitalized patients whose care was informed by an AI-based clinical deterioration early warning system (EWS) were 36% less likely to die during a pragmatic cluster-randomized trial when compared to patients on usual care units without the EWS (adjusted hazard ratio [HR], 0.64; 95% confidence interval [CI], 0.53– 0.78).^1^ The authors should be applauded for conducting a trial evaluating the effect of an AI intervention on patient health outcomes; more such trials are needed. Several aspects of the study, however, raise concerns that its findings may not accurately reflect the effect of the EWS. First, spillover effects on availability of scarce intensive care unit (ICU) beds could have led to worse outcomes for patients in the control group. Second, EWS-informed clinical decisions may have led to more frequent exclusion of high-risk patients from the intervention group. Either of these concerns could have led to overestimation of AI benefit. Finally, large pre-trial differences in hospital unit mortality rates suggest that unreported cluster-level imbalance or between-cluster variance could also have affected trial results.

This trial was conducted in 2 health systems in the United States, with each system containing one academic medical center and one community hospital.^1^ Individual hospital units (acute and intensive care) were randomized to implement the EWS or serve as a usual care control. The trial evaluated the COmmunicating Narrative Concerns Entered by RNs (CONCERN) EWS intervention, a machine learning algorithm that uses nursing surveillance documentation to identify patients at risk of clinical deterioration in the next 24 hours. For each monitored patient, the EWS presented a green (low), yellow (increased), or red (high) risk score to the care team with the goal of enabling early care escalation.

Our first concern is the potential for negative spillover due to constraints on care delivery.^2-4^ This occurs when a study intervention increases utilization of scarce medical resources and thus negatively affects care availability and outcomes for control group patients. Spillover can inflate an intervention’s apparent benefit during a trial, and may help explain why some interventions, when adopted at scale, do not improve outcomes as expected.

During the CONCERN-EWS trial, intervention group patients were 25% more likely to be transferred to the intensive care unit (ICU) than control group patients, suggesting that the EWS prompted care escalation as intended.^1^ Yet increased ICU utilization by the intervention group could also have led to reduced ICU bed availability for control group patients in the same hospital, particularly given pandemic-induced “poor staffing, increased patient acuity, and limited hospital resources” reported by the authors during the trial’s study period (2020-2022).^1^ Other studies have found patients not admitted to the ICU due to lack of beds face higher mortality;^5^ a post-hoc analysis reported that delayed ICU transfer was associated with higher mortality for a subset of patients with sepsis in the CONCERN-EWS trial.^6^

A second concern is that exclusion from the mortality analysis may have affected trial arms differently. Patient deaths were excluded if they occurred after initiation of palliative or hospice care, after do not resuscitate (DNR) or do not intubate (DNI) order activation, or after another outcome (cardiopulmonary arrest, sepsis, unanticipated ICU transfer) occurred.^1^ Following these exclusions, mortality rates reported during the trial (10.0 deaths per 10,000 inpatient days for the intervention group and 15.8 deaths per 10,000 inpatient days for the control group) were less than half the rate observed at study sites before the trial (37.5 deaths per 10,000 inpatient days).

The authors acknowledge that exclusions applied to patients with known high rates of mortality. They do not address the potential for differential exclusion across the intervention and control groups, however. Small differences across groups are apparent in the case of the one exclusion analyzed separately: in a subsequent publication, the trial investigators reported that 98 patients died in the hospital after unanticipated ICU transfer from intervention units and 107 died after transfer from control units.^7^ These deaths were not included in the trial results; had they been included and attributed based on group assignment of the originating unit, the reported number of deaths would be 54% higher (279; up from 181) in the intervention group and 40% higher (372; up from 265) in the control group.

Differences could be greater in the case of other exclusions, particularly if the EWS itself, by identifying patients at high risk of deterioration, influenced whether a patient’s death was excluded. Presented with a red alert in a patient’s EHR display, clinicians might be more likely to initiate a goals-of-care discussion that leads to a DNR/DNI order or palliative/hospice care and thus exclusion of subsequent death from the trial results. No such alert would inform care for a similar at-risk patient in the usual care control group; their death might instead be included in results. In other hospitals, AI systems that identify patients at risk of deterioration have led to increased rates of goals-of-care discussion, palliative care consultation, and DNR status.^8,9^ Outcomes for 1,499 patients with DNR/DNI, palliative, or hospice care status were excluded from the trial results; by comparison, 446 deaths were included. If differential exclusion of patient deaths occurred as a result of EWS-informed care decisions, this could sway the primary mortality findings.

A final set of concerns relates to whether outcomes differed across hospital units and how this was handled in statistical models. Our stratified analysis of the released de-identified trial data shows that all significant findings from the trial are reproduced at Site B where the EWS model was trained but not at Site A (Table 1).^10^ This raises questions about EWS effectiveness in other settings. Separately, differences in mortality across hospitals raise the possibility that chance imbalances between randomized groups affected the findings.

**Table 1.**
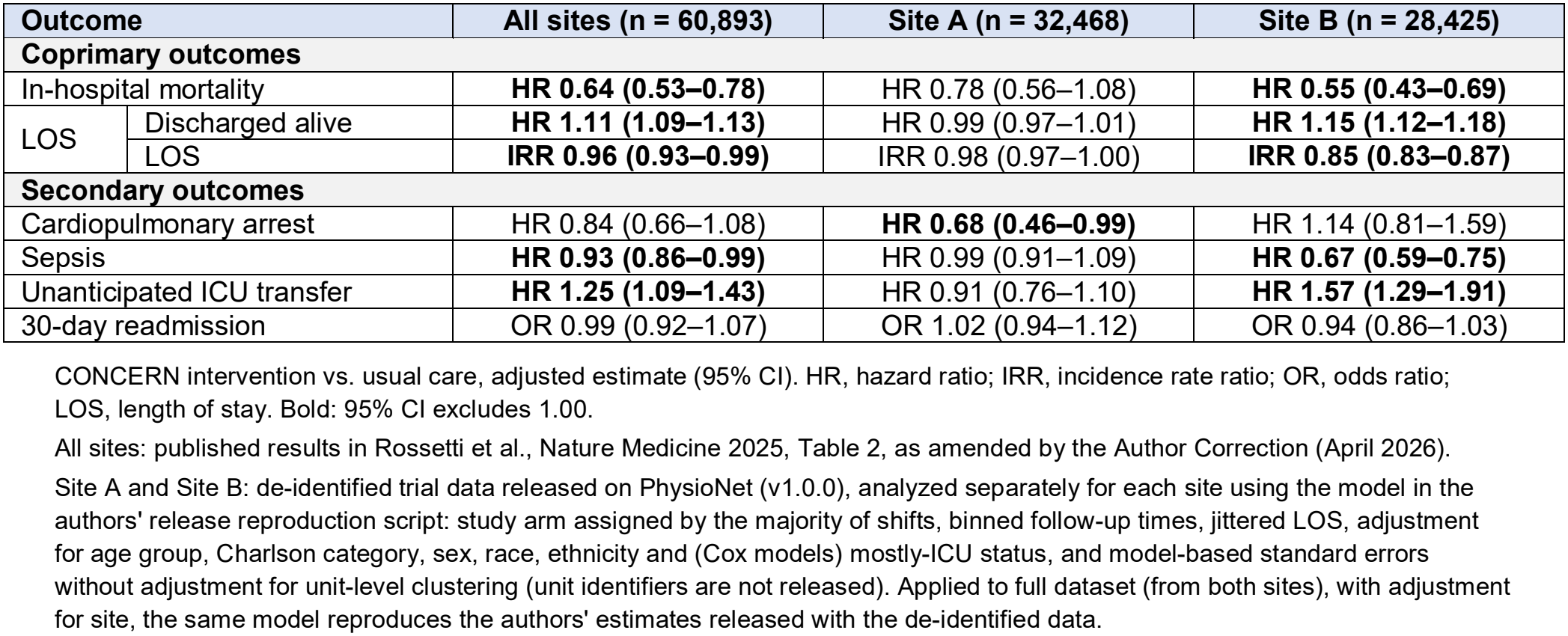
Adjusted effect estimates: published results for all sites compared to site-stratified analysis of released deidentified trial data.

At one site, mortality rates in 2014 were 11.9 deaths per 10,000 inpatient days in the 5-unit community hospital and 48.8 deaths per 10,000 inpatient days in the 21-unit academic medical center.^11,12^ Given this four-fold difference in pre-trial mortality rates, if 4 or more units in the low-mortality hospital were randomly assigned to one group, this alone could lead to an imbalance, at least at this one site. Within-hospital differences in unit mortality rates could further increase the probability of imbalance. No baseline unit-level characteristics are reported to allow comparison of intervention and control groups, however. Any unit-level imbalance in patient acuity or case mix resulting in higher mortality would not be detected by the authors’ assessment of patient-level balance using Charlson Comorbidity Index (CCI) scores as a proxy for illness severity; in analysis of de-identified trial data, higher CCI scores were not associated with mortality (HR 0.96 per point, 0.91-1.01).

The authors state that all analyses account for clustering at the hospital unit level, but they have not shared the SAS code used or reported the estimated unit-level frailty variance or other measure of clustering. The authors’ validation analysis of deidentified trial data does not account for clustering but produces confidence intervals of similar width as in the published cluster-adjusted results for all outcomes except corrected length of stay (Figure 1). Similar intervals do not necessarily indicate a problem, but the comparison raises questions about how clustering affected precision, particularly given large pre-trial differences in mortality between hospitals.

**Figure 1.**
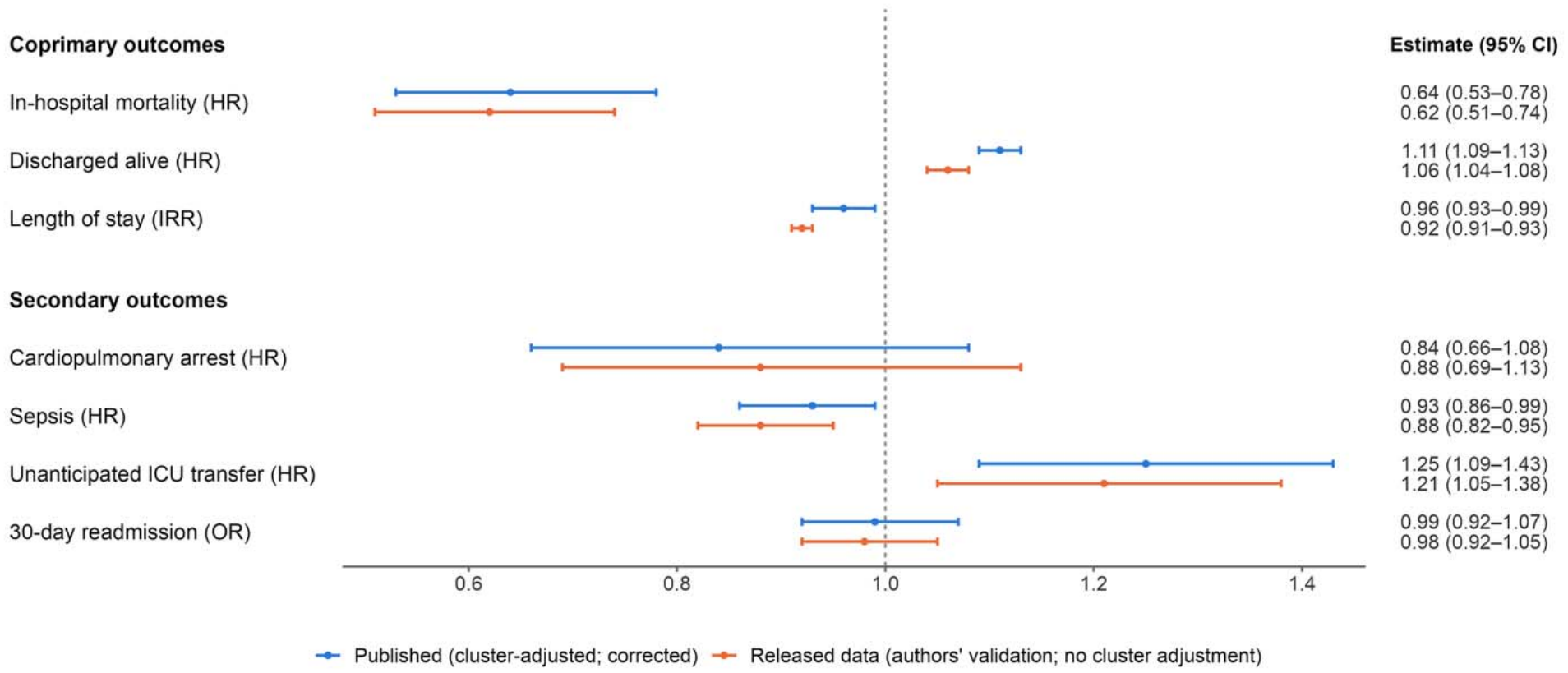
Published cluster-adjusted estimates compared with the authors’ analysis of released trial data without cluster adjustment. CONCERN intervention vs usual care, adjusted estimate and 95% confidence interval. HR, hazard ratio; IRR, incidence rate ratio; OR, odds ratio. Values below 1 indicate a lower rate or odds in the intervention group. **Published:** Published analyses accounted for unit-level clustering (frailty models for survival outcomes; generalized estimating equations for length of stay). From Rossetti et al., Nature Medicine 2025, Table 2, as amended by the Author Correction (April 2026), which revised the length of stay and 30-day readmission results (pre-correction estimates: length of stay IRR 0.91 [0.90–0.93]; 30-day readmission OR 0.99 [0.93–1.05]).^15^ **Released data:**estimates reported by the trial authors in the PhysioNet data release (v1.0.0) to validate the de-identified trial data, from models producing standard errors without accounting for unit-level clustering. Study arm assigned by the majority of shifts; binned follow-up times; jittered length of stay; adjusted for age group, Charlson category, sex, race, ethnicity, site and (Cox models) mostly-ICU status. Published confidence intervals that account for clustering are between 0.9 and 1.3 times as wide as the validation analysis confidence intervals without clustering, except for corrected length of stay which is over twice the width.

These concerns (apart from spillover) could be investigated using data available to the study authors. We encourage them to report baseline unit-level characteristics by study arm (including pre-trial mortality rates), the number of excluded admissions for each arm that resulted in death or discharge to hospice, and unit-level frailty variance or another measure of how mortality varied between hospital units. Much of this information is requested in the CONSORT extension for cluster-randomized trials (items 13b, 15, 17a),^13^ but the original study does not include it. Regardless of whether this additional information strengthens or weakens the trial’s main findings, it would be a valuable aid in interpreting trial results.

More broadly, future trials evaluating clinical AI will often confront similar concerns. AI tools that relieve or increase strain on shared healthcare resources raise the prospect of spillover. AI-prompted changes in care pathways, when combined with complex exclusion rules, can lead to differential censoring of outcomes. Cluster randomization to avoid contamination brings a higher risk of chance imbalance and requires careful cluster adjustment in statistical models. Transparent reporting is a key part of addressing these concerns.

## Methods

### Data source

We used the de-identified encounter-level dataset released by the trial investigators on PhysioNet (CONCERN Early Warning System Trial, version 1.0.0), accessed under the PhysioNet Restricted Health Data Use Agreement 1.5.0.^1,10,14^

The release contains one row for each of the 60,893 patient encounters (admissions) included in the trial analyses. The release provides study site (coded 1 = Site B and 2 = Site A in the release documentation); age group; sex; race; ethnicity; Charlson Comorbidity Index category (0, 1, 2, 3, 4, 5+); study arm, assigned as the arm of the units on which the encounter spent the majority of its shifts; an indicator for encounters spent mostly in intensive care; indicators for each outcome; follow-up for each survival outcome, binned into intervals of 1–2, 3–6, 7–14, 15–28 and 29 or more 12-hour shifts; and length of stay rounded and perturbed with random jitter. Unit identifiers are not included in the release.

### Statistical analysis

We used the model specification in the authors’ validation script released with the data. As described in the release documentation, in-hospital mortality, discharge alive, cardiopulmonary arrest, sepsis and unanticipated ICU transfer were analyzed with Cox proportional hazards models using the binned follow-up times, length of stay with negative binomial regression, and 30-day readmission among encounters discharged alive with logistic regression. The models were adjusted for age, sex, race, ethnicity, ICU status, Charlson Comorbidity Index category and study site. Applied to the full dataset, these models reproduce the estimates reported by the investigators in the release documentation (for example, mortality HR 0.62, 95% CI 0.51–0.74).

We conducted two additional analyses. First, site-stratified estimates (Table 1) were obtained by fitting the same models separately to encounters at Site A and Site B, omitting the site covariate. Second, to examine the association between comorbidity and mortality, the mortality model was refitted with the Charlson category replaced by a linear score from 0 to 5 (with 5+ scored as 5), retaining all other covariates, including study arm and site. The hazard ratio per point is reported for all encounters.

Analyses were performed in R 4.5.1 using the survival (3.8.3) and MASS (7.3.65) packages.

### Limitations

Because unit identifiers are not released, standard errors are model-based and do not account for clustering by unit; confidence intervals may be different than they would be under a cluster-adjusted analysis. The released data on PhysioNet also differ from those used in the published analyses: arm assignment is fixed at the encounter level rather than time-dependent, follow-up times are binned, and length of stay is jittered.

## Data Availability

The deidentified data used in our analysis are available on Physionet at https://physionet.org/content/concern-early-warning/1.0.0/.

## Author contribution statement

S.M. wrote the first draft of this paper and analyzed the data. All authors read, revised, and approved the final version.

## Ethics statement

This study used deidentified data from a public repository and was deemed exempt by RAND’s institutional review board (IRB Study ID: 2026-N0544).

## Data availability statement

The deidentified data used in our analysis are available on PhysioNet at https://physionet.org/content/concern-early-warning/1.0.0/.

## Code availability statement

The code used in our analysis is available at: Mann, S. (2026). Code to accompany ‘Concerns over spillover, exclusions, and hospital unit differences in an AI early warning system trial’ [Computer software]. Zenodo. https://doi.org/10.5281/zenodo.24477753

## AI use declaration

An LLM (Claude Opus 5.5) was used to draft R code for use in analysis and figure generation. Sean Mann reviewed all LLM outputs and takes responsibility for the integrity of the content generated.

